# SARS-CoV-2 Antibody response to the Sputnik Vaccine in previous infected Patients and non-infected one

**DOI:** 10.1101/2022.11.30.22282668

**Authors:** Mohamed Abdulsammad, Fawzi Ebrahim, Salah Tabal, Sedigh Bashir, Ashraf Aburgiga, Mohamed Milad, Mohamed Bareem

## Abstract

At the begging of 2020 saw the development and trials of vaccines against Covid-19 at an unprecedented pace. The first half of 2021 has seen vaccine rollout in many countries, on the other hand, Immunity to covid-19 has exhibited to minimize the risk of having a severe infection and initiate an excellent degree against the disease. This study focuses on the comparison of Anti-Spike IgG antibodies among vaccinated people with or without previous exposure to the coronavirus. To determine whether a single dose of sputnik V can produce significant antibody titer amongst previously infected cases and design vaccine dosage regimens accordingly. This study was performed at Libyan biotechnology research Centre from August 2021 to December 2021. Blood samples were collected from 1811 adult males and females vaccinated with and without a history of exposure to covid-19. Previously infected individuals’ record was noted separately. Samples were immediately analyzed by Beckman Unicel Dxl 600, Access immunoassay system. Data were analyzed using GraphPad Prism 9 Software. A P-value >0.5 was not significant. The Majority of candidates 60% of the total samples were males and on analysis, it was found that 72% of patients were seropositive, on the other hand, individuals who vaccinated and have naïve antibodies from the previous infection showed slightly higher immunological response rather than vaccinated patients without previous infection and this finding can help the policymakers to design a single-dose vaccine regimen for the former category.

## Introduction

The outbreak of COVID-19 has led to one of the most advanced vaccination programs ever. Immunity to SARS-CoV-2 either stimulated via natural infection or vaccination has been shown to minimize the risk of having a severe infection and initiate a good degree of protection against getting re-infected (Khoury *et al*., 2021). Seropositive individuals against SARS-CoV-2 have ensured reasonable protection of more than 80% from reinfection (Lumley *et al*., 2021). On the other hand, around 60-95% efficacy of vaccines have been reported and worldwide countries are competing to vaccinate their citizens against severe acute respiratory syndrome coronavirus 2 (Kim *et al*., 2021). Vaccines remain unclear and the only evidence available is the presence of primary immune responses (Gaebler *et al*., 2021). Another concern is the emergence of viral mutations that may be resistant to both vaccines induced and naïve immune responses (Wang *et al*., 2021). In Libya, the vaccine drive started early in 2021 with more than one million people fully vaccinated until mid of 2022 (World Health Organization, 2022). Libya has so far approved five Covid-19 vaccines; Sinopharm, Sinovac, Sputnik, AstraZeneca, and Pfizer. Phase 3 trials of these vaccines showed reasonable efficacy at preventing severe infection after two doses (or one if single dose vaccine) administered at a fixed time interval. A summary of these vaccines is given in table 1.

**Table 1:** Vaccines along with their mechanism of action.

| Company | Type ofVaccine | Mechanism |
| --- | --- | --- |
| Sinopharm | Inactivate Viral Vaccine | Virus killed byheat / chemicals |
| Sinovac | Inactivated Viral Vaccine | Virus killed byheat / chemicals |
| Sputnik | Viral Vectorvaccine | A harmless Virus transports Virus gene |
| AstraZeneca | Viral Vectorvaccine | A harmless Virus transports Virus gene |
| Pfizer | RNA Vaccine | mRNA template forViral proteins |

The vaccination drives through the vaccination center distributed entire Libya, started in June 2021 and one dose of Sputnik V was administered intramuscularly. Sputnik V is an adenovirus viral vector vaccine developed by Gamaleya Research Institute registered on 11 August 2020 by the Russian Government. Clinical trials have shown a strong protective immunological response to Sputnik V across all age groups. This vaccine uses adenovirus 26(Ad26) and adenovirus 5(Ad5) as vectors for the expression of the SARS-CoV-2 spike protein, and an efficacy efficacy of 91.6% was reported after the first dose of Sputnik V in a Russian Study and evidence of reduced disease severity is encouraging for the supporters of dose sparing strategy (Logunov *et al*., 2021). In a developing country such as Libya, with limited resources, studies conducted to look for the antibody titer after vaccination or previously infected individuals can be very helpful to assess the efficacy of vaccines and the immunological response after the first shot. This will help to design the dosage regimens accordingly. The persistence of vaccine-induced antibody levels is still not known, however, infection-induced antibodies tend to remain detectable almost 6 months after infection (Jones and Roy, 2021). Studies show that individuals who have been infected with the coronavirus can safely skip the second jab of two doses of vaccines, however, the concern lingers (Pradenas *et al*., 2021). Ideal vaccine dosage regimens have not yet been designed separately for those who have been naturally infected and those who have not been exposed to the virus or are antibody naïve. We designed this study to see whether vaccinated pre-infected individuals had a better immunological response as compared to uninfected individuals. This study can also help us to assess whether previously infected individuals could establish recall responses to a single shot of vaccine and the un-administered doses can be saved for the rest of the population.

## Materials and Methods

This study was conducted at Libyan biotechnology research center from August 2021 to December 2021. Ethical approval was obtained from Bioethics Committee at the Biotechnology Research Center in Tripoli under letter number BEC-BTRC 8-2020. Blood samples were collected from 1811 adult male and female patients who booked for the vaccine shot. Patients were not charged for the vaccination and antibody test. The samples were coded, and serum was separated and stored at −20°C until analyzed within 48 hours. The study protocol was compatible with the World Medical Association Declaration of Helsinki (Ethical Principles for Medical Research Involving Human Subjects). All participants provided written informed consent to participate. Those who agreed to participate were given an information sheet detailing the study’s aim, pledging anonymity of their information, and explaining that they have the right to withdraw from the study at any time. A self-administered questionnaire was used to collect data on sex, age, the type of vaccines received, the date(s) of vaccination, side effects, the severity of symptoms, previous Covid-19 infection, and whether the infection (if there was one) was before or after receiving the vaccine. Information on past medical history and influenza vaccination status was also noted.

### Detection of SARS-CoV-2 specific serum antibodies

The Beckman Coulter Access Anti-SARS-CoV-2 IgG assay was used on a UniCel Dxl 600 Access Immunoassay System to detect anti-SARS-CoV-2 antibodies according to the manufacturer’s instructions (Beckman Coulter, Germany. A sample was considered reactive (seropositive) for anti-S IgG if the result was ≥ 10 AU/ml.

## Results

Our study group comprised of 1811 candidates; 60% (n=1088) were males and 40% (n= 723) were females. A total of 1319 patients were RT-PCR confirmed SARS-CoV-2 positive at least 30-60 days after the first dose of the Sputnik vaccine based on the history they gave. On analysis, it was found that 73% of the patients were seropositive whereas 27% of the patients were seronegative, despite, all individuals receiving the first dose of the vaccine (table-2). Data were analyzed using GraphPad Prism 9 Software. Percentages were calculated and mean antibody titer between groups was compared using a t-test. p-value > 0. 5 was considered not significant.

The percentage of seropositive patients who did not expose to covid-19 and after getting the first shot of the vaccine was more minor as compared to those who gave a history of being infected and getting the first dose of Sputnik V (Table 2). The mean age of seropositive patients was 35.4 a year, and the mean antibody titers of vaccinated participants with the previous infection was 115 AU/ml versus 102 AU\ml for vaccinated patients who did not expose to SARAS-COV-2 according to data shown in table-3. We finally divided the study participants on seropositivity and compared the mean antibody titer between seropositive study participants who get one shot of the sputnik vaccine with pre-infection versus individuals without previous infection of covid-19. It was found that a slight difference was present in the antibody titers of the two groups (Table 3).

**Table 2:**
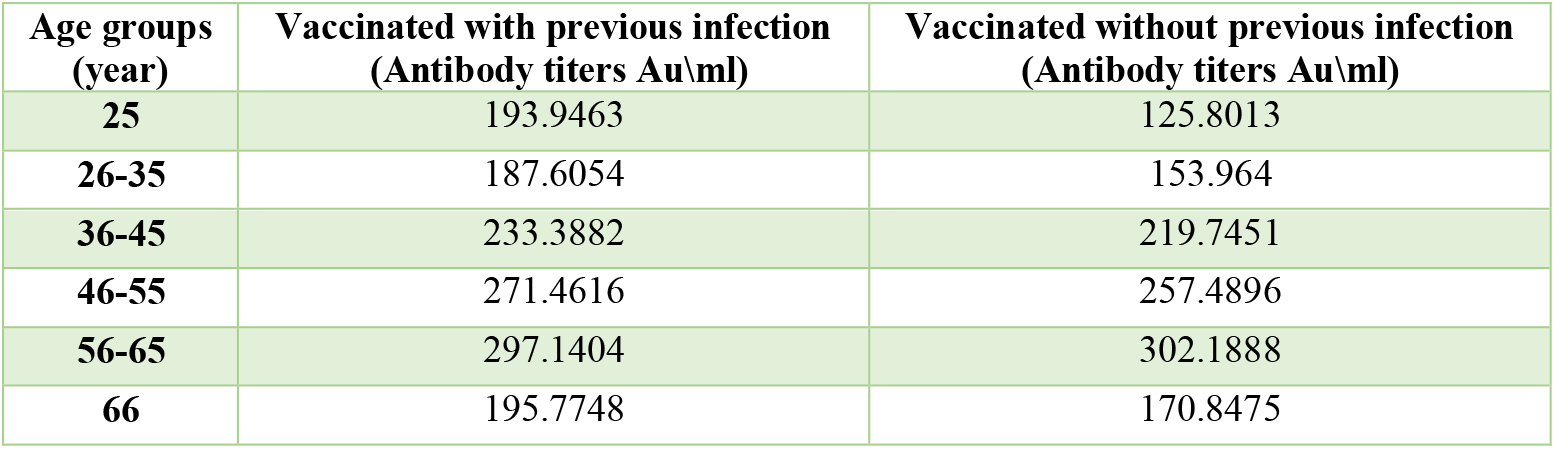
The difference in percentage, gender, and reactivity of vaccinated patients with or without covid-19 previous infection.

| Age groups (year) | Vaccinated with previous infection (Antibody titers Au\ml) | Vaccinated without previous infection (Antibody titers Au\ml) |
| --- | --- | --- |
| 25 | 193.9463 | 125.8013 |
| 26-35 | 187.6054 | 153.964 |
| 36-45 | 233.3882 | 219.7451 |
| 46-55 | 271.4616 | 257.4896 |
| 56-65 | 297.1404 | 302.1888 |
| 66 | 195.7748 | 170.8475 |

**Table 3:** comparison between antibody titers of vaccinated patients with or without previous infection according to age groups.

| Participant | Total | M | % | F | % | R | M | % | F | % | Non R | M | % | F | % |
| --- | --- | --- | --- | --- | --- | --- | --- | --- | --- | --- | --- | --- | --- | --- | --- |
| vaccinated with Pre. Infection | 1537 | 893 | 58 | 644 | 42 | 1144 | 631 | 55 | 513 | 45 | 393 | 262 | 66 | 131 | 33 |
| vaccinated without Pre. Infection | 274 | 195 | 71 | 79 | 29 | 175 | 132 | 75 | 43 | 25 | 99 | 63 | 63 | 36 | 36 |
| Total | 1811 | 1088 | 60 | 723 | 40 | 1319 | 763 | 57 | 556 | 42 | 492 | 325 | 66 | 167 | 33 |

M=male, F=female, R=reactive =seropositive.

## Discussion

Coronavirus emergence accelerate the invention, development, and production of nine vaccines based on different production techniques. The Government of Libya started this vaccination drive after the vaccines were confirmed safe to use by the World Health Organization. The Libyan National Center for Disease Control initiated this drive to fulfill the huge requirement of the population getting vaccinated in the public sector. Public health institutes enabled us to start an independent follow-up study of vaccination-induced immunity. In the present study, we observed a little difference in the immune response of vaccinated candidates without previous covid-19 infection and another individual who was vaccinated with a history of the disease, both groups responded to the first dose of vaccine producing detectable antibodies but the percentage of seropositive participants who did not expose to covid-19 was more minor as compared to those who gave a history of being infected and getting the first dose of Sputnik vaccine. The vaccine-induced immune response is found to be higher in patients having severe disease courses as compared to those having asymptomatic disease according to latest studies (Virtanen *et al*., 2021; Lynch *et al*., 2020; Grossberg *et al*., 2021). In this study, we did not evaluate the patients according to the disease severity, therefore we couldn’t comment on the correlation of the severity of infection with the antibody titer. However, there was a considerable increase in the titer after the second dose which is in accordance with the study performed by (Jalkanen *et al*., 2021). The authors of the aforementioned study suggested a single-shot vaccination strategy or low-priority stratification for previously infected individuals due to the worldwide vaccine shortage (HAS 2021). A recent study showed findings nearly similar like to our study; Previously infected study participants had slightly higher titers of the anti-spike antibody as compared to those without history of infection. The authors of this study also mentioned that a single vaccine shot was enough to develop a significant vaccine titer in the seropositive group (Bradly *et al*., 2021). Lombardi et al reviewed antibody titers among vaccinated healthcare employees and stratified the results on the basis of the previous history of infection. The researchers found that the health care workers who were infected more than 6 months before getting the vaccine had significantly higher titers of the antibody as compared to those who were infected less than 6 months before vaccination and the uninfected study subjects had the lowest titers of antibodies (Lombardi *et al*., 2021). Higher antibody titers among previously infected candidates are an expected finding as the vaccine acts as a booster of natural immunity. Many studies show similar findings and some of the reasons given by researchers include multiple exposures to SARS-CoV-2 acting as natural boosters and vaccination at longer time intervals after getting naturally infected leading to higher anamnestic response (Manisty *et al*., 2021; Krammer *et al*., 2021; Ebinger *et al*., 2021; Anichini *et al*., 2021). Researchers have also proven that single-dose vaccination is able to elicit an anamnestic response in seropositive individuals and these antibodies are capable of neutralizing heterologous antigens effectively (Purushotham *et al*., 2021). Owing to the scarce vaccine supply and financial constraints in the underdeveloped world, many studies were conducted to see whether a single dose is enough for pre-infected individuals or not. Many of those studies showed that the second jab of a two-dose vaccine regimen can be easily skipped for the pre-infected cases owing to high antibody titers after a single dose. Our study also revealed nearly similar findings and it can be suggested that one dose regimen can be used for seropositive individuals, however, more evidence is still required.

## Conclusion

In summary our study showed seropositive vaccines with a history of pre-infection to covid-19 have a slightly elevated antibody titer as compared to seropositive subjects who did not expose to SARS-CoV-2 infection. These findings can be used by health policymakers to develop a single-dose vaccine regimen for selected populations and better use of health finances in a developing country like Libya.

## Data Availability

All data produced are available online at
https://docs.google.com/spreadsheets/d/18ozxwOwfzSu984AWimJvblniJSfrnm-7/edit#gid=1414940867

https://docs.google.com/spreadsheets/d/18ozxwOwfzSu984AWimJvblniJSfrnm-7/edit?usp=sharing&ouid=115066519336643469724&rtpof=true&sd=true

## Notes

### Competing Interest Statement

The authors have declared no competing interest.

### Clinical Protocols

https://www.wma.net/policies-post/wma-declaration-of-helsinki-ethical-principles-for-medical-research-involving-human-subjects/

### Funding Statement

This study did not receive any funding.

### Author Declarations

Ethical approval was obtained from Bioethics Committee at the Biotechnology Research Center in Tripoli under letter number BEC-BTRC 8-2020.

